# Prevalence of diarrheagenic *Escherichia coli* and impact on child health in Cap-Haitien, Haiti

**DOI:** 10.1101/2022.10.10.22280899

**Authors:** Jenna N. Diaz, Lora L. Iannotti, Sherlie Jean Louis Dulience, Sydney Vie, Xuntian Jiang, Vadim Grigura, Jacques Boncy, Francesca J. Marhône Pierre, F. Matthew Kuhlmann

## Abstract

**Background:** Diarrheagenic Escherichia *coli* (DEC) are common pathogens infecting children during their growth and development. Determining the epidemiology and the impact of DEC on child anthropometric measures informs prioritization of prevention efforts. These relationships were evaluated in a novel setting, Cap-Haitien, Haiti.

**Methods:** A case-control study of children 6-36 months of age enrolled 99 cases with diarrhea and 96 asymptomatic controls. Assessments were performed at enrollment and one month follow-up. Established PCR methodologies targeted DEC using fecal swabs. The association between pathogens and anthropometric z-scores was determined using multiple linear regression.

**Results:** Enterotoxigenic *Escherichia coli* (ETEC) was identified in 21.9% of cases vs. 16.1% of controls with heat-stable producing ETEC significantly associated with symptomatic disease. Enteroaggregative *E. coli* (EAEC) was found in 30.2% of cases vs. 27.3% of controls, and typical enteropathogenic *E. coli* in 6.3% vs. 4.0% of cases and controls, respectively. Multivariate linear regression demonstrated ETEC and EAEC were significantly associated with reduced weight-age z-score (WAZ) and height-age z-score (HAZ) after adjusting for confounders.

**Conclusions:** DEC are prevalent in north Haitian children. ETEC, EAEC, household environment, and diet are associated with unfavorable anthropometric measures. Further studies may quantify the contribution of individual pathogens to adverse health outcomes.

**Author Summary:** Multiple factors contribute to poor child growth and development including infectious diarrhea, malnutrition, and water, sanitation, and hygiene infrastructure. Efforts to improve child development require prioritization based on their cost and expected benefits. This study contributes to prioritization efforts by determining the local burden of disease in an understudied region, Cap-Haitien, Haiti, and associating this burden with measures of child growth. Moreover, it captures data on social determinants contributing to child growth to control for confounding variables. Unlike prior studies in Haiti, we evaluated children with diarrheal disease in the community instead of the hospital setting. Ultimately, we showed that Enterotoxigenic *Escherichia coli* and Enteroaggregative *E. coli* are common and associated with surrogate measures of poor child growth. This result suggests that ETEC vaccination efforts may have a strong effect on improving child health. This work supports the continued investments in the development of ETEC vaccines.

## Introduction

Mortality from diarrheal disease continues to decline more rapidly than disease incidence. As a result, complications from diarrheal disease persist (1, 2). Repeated episodes of diarrhea also increase risks of long-term complications, such as growth faltering and cognitive deficits (3, 4). Therefore, preventing this morbidity remains a public health priority. Multiple infectious agents cause diarrheal disease and their individual contributions to poor health outcomes remains challenging to quantify (5, 6). Effective prevention strategies such as water, sanitation, and hygiene (WASH) infrastructure, behavior change, nutritional interventions, and vaccinations can contribute differentially to improved health.

Moreover, each strategy requires differing investments in time or financial resources to be successful. With limited resources and competing interests, quantifying the impact of distinct interventions relative to expected health outcomes helps prioritize distinct strategies (7).

Diarrheagenic *Escherichia coli* (DEC) describes several common, pathogenic *Escherichia coli (E. coli)* pathovars with diverse disease manifestations (8-10). Enterotoxigenic *E. coli* (ETEC) is one such pathovar, ranging in clinical severity from asymptomatic to severe watery diarrhea. ETEC expresses either the heat-stable toxin (ST), and/or the heat-labile toxin (LT). Early studies focused solely on ETEC demonstrate an association with impaired growth (11). Enteroaggregative *E. coli* (EAEC) is associated with less severe disease, but asymptomatic colonization may impair growth (12). They are defined phenotypically by forming “stacked bricks” aggregates on contact with intestinal epithelia. Despite this clear phenotype, the molecular characterization of this pathovar remains unclear (13). Finally, enteropathogenic *E. coli* (EPEC) are less common and present with a range of diarrheal severity. Typical EPEC (tEPEC) express both the attachment and effacing (eae) and the bundle forming pillus (bfpa) genes (8). Their association with long-term complications is less clear.

Variations in local epidemiology, diet, and environment alter the impact of enteropathogens on health outcomes, making community-specific evaluations critical (14, 15). Also, extrapolating disease burden estimates to areas without detailed country-level information remains problematic (15, 16). In Haiti, high rates of diarrheal disease persist despite eliminating cholera and rotavirus through sustained vaccination efforts (17, 18). Nearly 40% of children under five years old experience diarrhea over a two-week interval, a prevalence that is unchanged during the past decade (19, 20). This high level of community-based disease persists while epidemiological studies have focused on medically attended diarrhea (17, 21-24). WASH strategies face significant hurdles in Haiti where 65% of households have access to clean water and 50% have unsatisfactory waste management systems (25). Local infrastructure lacks proper landfills and communities such as Cap-Haitien struggle with high population densities (26). Cap-Haitien is the second largest city in Haiti, located on the northern coast, and is distinct from the capital, Port-au-Prince, with lower crime rates but less development. Malnutrition combined with diarrheal disease further increases the risk of long-term disability in affected children and remains common in Haiti (27-29). Varied dietary patterns and nutrient deficiencies specific to Haitian children may influence the risk of diarrheal disease or adverse outcomes (30). The combination of diarrheal disease and malnutrition also induces long-term intestinal damage in the form of environmental enteropathy (EE) (4, 29). Therefore, these local contexts necessitate studies to identify effective, Haitian-specific prevention efforts (14-16).

Locally-available and accessible animal source foods (ASFs) may help reduce stunting. A randomized trial in Ecuador utilized eggs to significantly reduce childhood stunting but was associated with increased diarrheal disease (31). Two nutrients were inversely correlated with the growth outcome, docosahexaenoic acid (DHA) and choline (31, 32). DHA has anti-inflammatory properties and clinical trials evaluate its effects on child growth and development (33, 34). Choline influences neurocognitive development and deficiencies may induce intestinal inflammation similar to EE (34, 35). How these biomarkers may influence diarrheal disease outcomes remains unknown.

To address these concerns, we conducted a case-control study in Cap-Haitien, Haiti, with the following aims: 1) generate preliminary estimates of community-based DEC prevalence; 2) determine associations between DHA or choline levels and diarrheal disease; and 3) assess the association of specific pathogens with child anthropometry. Our pre-specified hypothesis stated that choline or DHA deficiencies are associated with more severe consequences of intestinal infection. Our additional hypothesis states that DEC are associated with poor anthropometric markers in Haitian children. This work demonstrates a high burden of DEC in community-dwelling, undernourished children in Cap-Haitien. Using the existing research infrastructure, additional studies to confirm these findings and estimate the impact of specific prevention efforts on outcomes will be performed (36).

## Methods

### Study design and participants

The longitudinal case-control study design was previously described (37). In brief, children aged 6-36 months and caregivers (>18 y.o.) living in Cap-Haitien, Haiti, were recruited from December, 2020 to May, 2021. Children with severe illnesses requiring emergent medical care were excluded, but this event did not occur. At enrollment, anthropometric measures, plasma samples, and fecal swabs were obtained on all children. In addition, enumerators administered a survey to caregivers, collecting information on socioeconomic status (SES), dietary behaviors, food intake frequencies, and WASH practices. These assessments were repeated one month later.

### Ethics statement

The Washington University Institutional Review Board (ID#202007027) and the Comité National de Bioéthique in Haiti approved the study. Written informed consent was obtained from adult caregivers in the native Creole language while children were too young to provide assent. All procedures were performed per the human experimentation guidelines of the United States Department of Health and Human Services and those set forth by Washington University for clinical research. Plasma samples were collected per the World Health Organization’s (WHO) guidelines (38).

### Group allocation

Cases were defined using a standard epidemiological definition (care-giver report of ≥ three liquid/semi-liquid stools in a 24 hour period over the preceding three days). Those without diarrhea at enrollment were defined as controls. Diarrheal symptoms were assessed again, one month later. Participants are subsequently defined as cases or controls, with or without diarrheal symptoms at follow up. Sample size for this pilot study was determined by logistical constraints to inform future trials.

### Data collection

#### Surveys

Trained nurses completed the surveys in the native Creole language. At baseline, demographic and SES information was obtained. In addition, children’s health, diarrheal history, and dietary intake were obtained at baseline and follow-up.

Dietary intake was assessed using a previously validated, 24-hour food frequency questionnaire (FFQ) (39). Minimum dietary diversity (MDD) and household dietary diversity score (HDDS) were then calculated based on WHO and Food and Agriculture Organization guidelines (40). For MDD, foods were categorized into specific groups: 1) grains, roots, and tubers; 2) legumes and nuts; 3) dairy products; 4) flesh foods; 5) eggs; 6) vitamin A-rich fruits and vegetables; 7) other fruits and vegetables: 8) breastmilk. HDDS groups included: 1) cereals; 2) roots and tubers; 3) vegetables; 4) fruits; 5) meat and poultry; 6) eggs; 7) fish and seafood; 8) legumes and nuts; 8) dairy products; 9) oil and fats; 10) sugar and honey; 11) miscellaneous. ASFs included any intake of red meat, chicken, eggs, seafood, or dairy products in the last 24 hours.

Health surveys included a 14-day morbidity recall, assessing the presence of fever, respiratory symptoms, rash, duration of diarrhea, presence of dysentery, or vomiting. Polio, rotavirus, and typhoid vaccination history was determined. Definitions for critical variables are provided in **Table S1**.

#### Anthropometry

Trained nursing staff used WHO protocols for standardized growth parameters to obtain anthropometric data using a Seca Model 874 digital scale for weight and a ShorrBoard® stadiometer for length. Measurements were taken twice, and differences greater than 0.1 kg for weight or 0.7 cm for length were repeated a third time. The two closest measurements were averaged. Children ≥ 2 years had 0.7 cm subtracted from their height to correct for recumbent measurements. The WHO Anthro Survey Analyser Software (v3.2.2) calculated height-for-age-z-score (HAZ), weight-for-age-z-score (WAZ), and weight-height-z-score (WHZ) (41). Children with z-scores of < -2 SD were considered stunted, underweight, and wasted, respectively.

#### Plasma and fecal samples

Trained phlebotomists collected plasma using lithium-heparin-containing BD Vacutainers^™^ at both timepoints according to WHO standards (38). Samples were transported on ice to the Hôpital Universitaire Justinien (HUJ) laboratory, centrifuged at ∼1200 x g for 20 minutes, aliquoted, and stored at -20°C. Caregivers obtained rectal swabs (Copan FecalSwab(tm) with Carey-Blair media) from each child at both visits unless requesting the trained nurses to collect the samples. Samples were transported from HUJ to the Kuhlmann laboratory at Washington University in St. Louis through the Haitian National Laboratory and stored long-term at -80°C.

### Plasma analysis

Plasma choline, DHA, and betaine concentrations were determined by modified liquid chromatography-tandem mass spectrometry (LC-MS/MS) as previously described (32) on a randomly selected subset of participants.

### Pathogen isolation and identification

Fecal swabs in Cary-Blair were vortexed with glass beads. Half the sample was used to extract RNA (EZ Tissue/Cell Total RNA Mini Kit, EZ BioResearch, R1002) according to the manufacturer’s protocol. gDNA was then extracted using the PureLink^™^ Microbiome DNA Purification Kit (Invitrogen, A29790). Eluted nucleic acids were stored at -80°C.

We targeted three commonly identified pathovars of DEC; ETEC, EPEC, and EAEC. Qualitative PCR amplification of unique molecular targets was performed using validated primers and tested against strains of known provenance (**Table S2**). The gene targets included: were *eltB, estA*, and *estB* for ETEC plus additional virulence factors, *eatA* and *etpA*; *eae* and *bfpA* for EPEC; *aaiC, aatA*, and a multiplex assay detecting four of five aggregative adherence fimbriae (aaf) for EAEC. ETEC was defined by the presence of ST (*estA* or *estB*) and/or LT (*eltB*) and grouped as ST or ST/LT ETEC compared with LT-only ETEC as in prior studies (16). Typical EPEC was determined by the presence of both *eae* and *bfpA*. EAEC was defined by the presence of both *aaiC* and *aatA* genes, with additional classification based on the presence of any of the four targeted aaf genes.

### Statistical analysis

The data utilized in this study is provided as supporting information (Supporting Information, Database). Descriptive statistics examined distributions for SES characteristics, the prevalence of *E. coli* pathovars, child anthropometry, and plasma concentrations of DHA, choline, or choline’s metabolite, betaine. Continuous variables were assessed for normal distribution and outliers using histograms, scatterplots, and boxplots.

Univariate analyses examined the differences in baseline characteristics for cases and controls. Continuous variables were assessed for significant differences using independent samples t-test. When equal variances were not assumed, variables were assessed using Welch’s t-test. Variables lacking normal distribution were assessed using Mann-Whitney U test. Categorical variables were assessed by chi-squared or Fisher’s exact test as appropriate. A dichotomous variable was created for stunting (HAZ < -2) and wasting (WAZ < -2).

Multivariate linear regression analysis was used to evaluate the association of ETEC and EAEC with anthropometric z-scores at baseline, adjusting for confounding factors relevant to the Haitian context. Factors considered for inclusion in the models included age, sex, household characteristics, WASH variables, and dietary intake variables. Backward stepwise regression determined which covariates were retained in final models. The interaction between ETEC and EAEC and the association of this interaction term with each anthropometric measure was tested. Covariates used in stepwise regression were added to the main effects of ETEC and EAEC, as well as the interaction effect. Diagnostics were evaluated to assess linearity, multicollinearity, homoscedasticity, and normality of residuals. All models met the assumptions of multivariate linear regression. For all analyses, the type I error was set to be two-sided and at 0.05. Analyses were completed using SPSS software (version 27.0) or R (version 4.1.3).

## Results

### Baseline characteristics relative to diarrheal disease

A total of 195 children (96 cases, 99 controls) completed baseline enrollment. Baseline demographics were previously reported, showing increased vomiting in cases over controls (37). Children completing follow-up (N=136) had reduced respiratory symptoms at baseline compared to those lost to follow-up (**Table S3**). Logistical and time constraints limited efforts to contact participants lost to follow-up.

We next evaluated baseline SES and demographic factors based on case-control status and follow-up symptoms **(Table 1, S4)**. Several findings were expected based on existing literature. First, cases with diarrheal symptoms at follow-up (column 4) were significantly more likely to have dirt or rock flooring and lower WHZ than the other groups. Cases without symptoms at follow up (column 3) had higher rates of vomiting, in keeping with a diagnosis of acute gastroenteritis at enrollment. Controls without symptoms at follow up (column 1) had non-significantly higher maternal education and older mothers, consistent with the assumption that higher SES is associated with lower rates of illness. As reported previously, cases had higher access to electricity than controls (columns 3 and 4 vs. 1 and 2) (37).

**Table 1.**
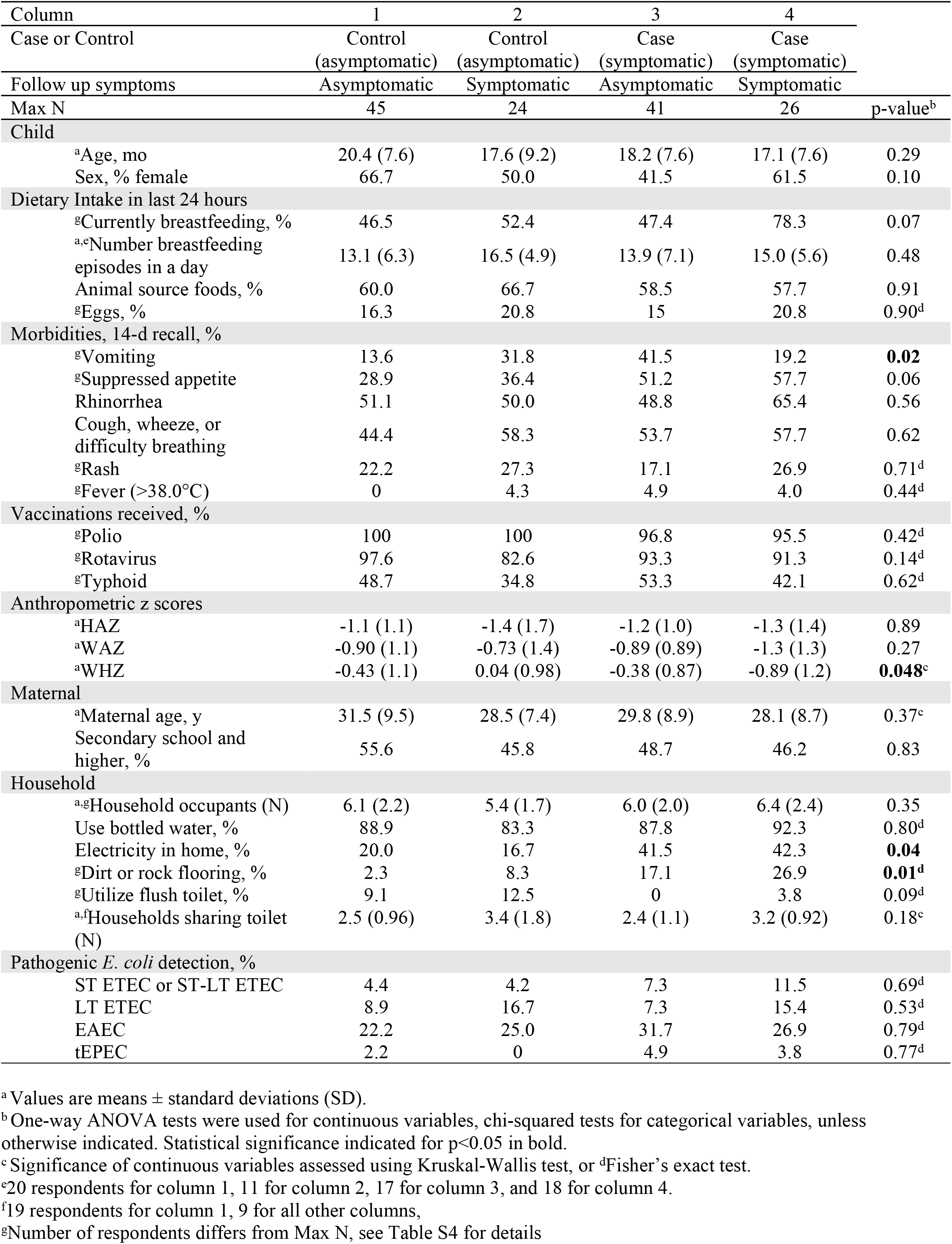
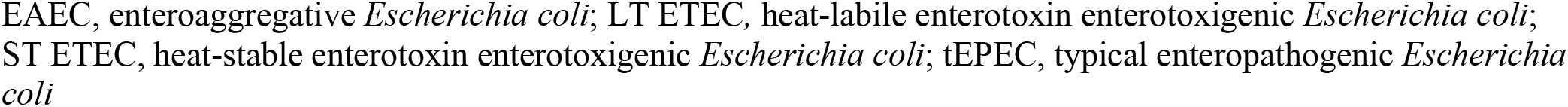
Association of baseline characteristics and diarrheal disease.

| Column | 1 | 2 | 3 | 4 |  |
| --- | --- | --- | --- | --- | --- |
| Case or Control | Control | Control | Case | Case |  |
|  | (asymptomatic) | (asymptomatic) | (symptomatic) | (symptomatic) |  |
| Follow up symptoms | Asymptomatic | Symptomatic | Asymptomatic | Symptomatic |  |
| Max N | 45 | 24 | 41 | 26 | p-value<sup>b</sup> |
| **Child** |  |  |  |  |  |
| <sup>a</sup>Age, mo | 20.4 (7.6) | 17.6 (9.2) | 18.2 (7.6) | 17.1 (7.6) | 0.29 |
| Sex, % female | 66.7 | 50.0 | 41.5 | 61.5 | 0.10 |
| **Dietary Intake in last 24 hours** |  |  |  |  |  |
| <sup>g</sup>Currently breastfeeding, % | 46.5 | 52.4 | 47.4 | 78.3 | 0.07 |
| <sup>a,e</sup>Number breastfeeding episodes in a day | 13.1 (6.3) | 16.5 (4.9) | 13.9 (7.1) | 15.0 (5.6) | 0.48 |
| Animal source foods, % | 60.0 | 66.7 | 58.5 | 57.7 | 0.91 |
| <sup>g</sup>Eggs, % | 16.3 | 20.8 | 15 | 20.8 | 0.90<sup>d</sup> |
| **Morbidities, 14-d recall, %** |  |  |  |  |  |
| <sup>g</sup>Vomiting | 13.6 | 31.8 | 41.5 | 19.2 | **0.02** |
| <sup>g</sup>Suppressed appetite | 28.9 | 36.4 | 51.2 | 57.7 | 0.06 |
| Rhinorrhea | 51.1 | 50.0 | 48.8 | 65.4 | 0.56 |
| Cough, wheeze, or difficulty breathing | 44.4 | 58.3 | 53.7 | 57.7 | 0.62 |
| <sup>g</sup>Rash | 22.2 | 27.3 | 17.1 | 26.9 | 0.71<sup>d</sup> |
| <sup>g</sup>Fever (>38.0°C) | 0 | 4.3 | 4.9 | 4.0 | 0.44<sup>d</sup> |
| **Vaccinations received, %** |  |  |  |  |  |
| <sup>g</sup>Polio | 100 | 100 | 96.8 | 95.5 | 0.42<sup>d</sup> |
| <sup>g</sup>Rotavirus | 97.6 | 82.6 | 93.3 | 91.3 | 0.14<sup>d</sup> |
| <sup>g</sup>Typhoid | 48.7 | 34.8 | 53.3 | 42.1 | 0.62<sup>d</sup> |
| **Anthropometric z scores** |  |  |  |  |  |
| <sup>a</sup>HAZ | -1.1 (1.1) | -1.4 (1.7) | -1.2 (1.0) | -1.3 (1.4) | 0.89 |
| <sup>a</sup>WAZ | -0.90 (1.1) | -0.73 (1.4) | -0.89 (0.89) | -1.3 (1.3) | 0.27 |
| <sup>a</sup>WHZ | -0.43 (1.1) | 0.04 (0.98) | -0.38 (0.87) | -0.89 (1.2) | **0.048<sup>c</sup>** |
| **Maternal** |  |  |  |  |  |
| <sup>a</sup>Maternal age, y | 31.5 (9.5) | 28.5 (7.4) | 29.8 (8.9) | 28.1 (8.7) | 0.37<sup>c</sup> |
| Secondary school and higher, % | 55.6 | 45.8 | 48.7 | 46.2 | 0.83 |
| **Household** |  |  |  |  |  |
| <sup>a,g</sup>Household occupants (N) | 6.1 (2.2) | 5.4 (1.7) | 6.0 (2.0) | 6.4 (2.4) | 0.35 |
| Use bottled water, % | 88.9 | 83.3 | 87.8 | 92.3 | 0.80<sup>d</sup> |
| Electricity in home, % | 20.0 | 16.7 | 41.5 | 42.3 | **0.04** |
| <sup>g</sup>Dirt or rock flooring, % | 2.3 | 8.3 | 17.1 | 26.9 | **0.01<sup>d</sup>** |
| <sup>g</sup>Utilize flush toilet, % | 9.1 | 12.5 | 0 | 3.8 | 0.09<sup>d</sup> |
| <sup>a,f</sup>Households sharing toilet (N) | 2.5 (0.96) | 3.4 (1.8) | 2.4 (1.1) | 3.2 (0.92) | 0.18<sup>c</sup> |
| **Pathogenic *E. coli* detection, %** |  |  |  |  |  |
| ST ETEC or ST-LT ETEC | 4.4 | 4.2 | 7.3 | 11.5 | 0.69<sup>d</sup> |
| LT ETEC | 8.9 | 16.7 | 7.3 | 15.4 | 0.53<sup>d</sup> |
| EAEC | 22.2 | 25.0 | 31.7 | 26.9 | 0.79<sup>d</sup> |
| tEPEC | 2.2 | 0 | 4.9 | 3.8 | 0.77<sup>d</sup> |

<sup>a</sup> Values are means ± standard deviations (SD).
<sup>b</sup> One-way ANOVA tests were used for continuous variables, chi-squared tests for categorical variables, unless otherwise indicated. Statistical significance indicated for p<0.05 in bold.
<sup>c</sup> Significance of continuous variables assessed using Kruskal-Wallis test, or <sup>d</sup>Fisher's exact test.
<sup>e</sup>20 respondents for column 1, 11 for column 2, 17 for column 3, and 18 for column 4.
<sup>f</sup>19 respondents for column 1, 9 for all other columns,
<sup>g</sup>Number of respondents differs from Max N, see Table S4 for details
EAEC, enteroaggregative *Escherichia coli*; LT ETEC, heat-labile enterotoxin enterotoxigenic *Escherichia coli*; ST ETEC, heat-stable enterotoxin enterotoxigenic *Escherichia coli*; tEPEC, typical enteropathogenic *Escherichia* *coli*

### Diarrheagenic Escherichia coli prevalence and associations with diarrheal disease

We evaluated the association of DEC with community-based diarrheal disease. Regarding ETEC, 21.9% of cases and 16.1% of controls carried ETEC (**Table 2**). Those with ST or ST/LT ETEC were more likely to be cases. Neither ETEC virulence factor, EatA or EtpA, were associated with cases but EatA was found more commonly in cases versus controls (9 vs. 4), consistent with prior work in Bangladesh (42). No differences between cases and controls were observed for EPEC. There was no difference in mean age for cases or controls with ETEC (p=0.77) or EPEC (p=0.66, Mann-Whitney U).

**Table 2.** Percentage of children with pathogenic *E. coli* by symptoms at both time points.

| Pathogen | Baseline |  | Total | p-value<sup>a</sup> | Follow-up |  | Total | p-value<sup>a</sup> |
| --- | --- | --- | --- | --- | --- | --- | --- | --- |
|  | Cases<br>(symptomatic)<br>(n=96) | Controls<br>(asymptomatic)<br>(n=99) |  |  | Symptomatic<br>(n=48) | Asymptomatic<br>(n=84) |  |  |
| ST or<br>ST-LT ETEC | 14.6 | 4.0 | 9.2 | **0.017** | 4.2 | 4.8 | 4.5 | 0.875 |
| LT ETEC | 7.3 | 12.1 | 9.7 | 0.350 | 12.5 | 15.5 | 14.4 | 0.639 |
| EAEC | 30.2 | 27.3 | 28.7 | 0.651 | 22.9 | 32.1 | 28.8 | 0.260 |
| tEPEC | 6.3 | 4.0 | 5.1 | 0.484 | 4.2 | 2.4 | 3.0 | 0.565 |

<sup>a</sup> Determined by chi-square testing.
EAEC, enteroaggregative *Escherichia coli*; LT ETEC, heat-labile enterotoxin enterotoxigenic *Escherichia coli*; ST ETEC, heat-stable enterotoxin enterotoxigenic *Escherichia coli*; tEPEC, typical enteropathogenic *Escherichia coli*

The molecular classification of EAEC remains challenging, however, we evaluated commonly accepted genes to define EAEC and potential alternative genes associated with virulence. No significant differences were observed between cases and controls using *aata* and *aaic*. Aggregative associated fimbriae (*aaf*) may modulate virulence, but again, we noted no significant differences as only 5.2% of cases and 5.1% of controls had isolates carrying these genes at baseline (Chi-squared, p=0.96). At follow-up, aaf genes were present in 2.1% vs. 2.4% of those with or without symptoms, respectively (Chi-squared, p=0.904). Cases with EAEC were younger than those without EAEC (13.7 ± 7.0 vs.18.6 ± 7.3, mean months ± 1SD, Student’s t-test, p=0.003), controls with EAEC were also younger but the difference was not significant. Carriage of multiple DEC was common but similar between cases and controls (**Table S5**).

Seasonal variations are known to affect the risk of diarrheal disease with increased infections during rainy seasons. In Cap-Haitien, the rainy season (April-June) was associated with increased ETEC prevalence regardless of symptoms (**Table S6**). Overall, we confirm a high prevalence of these pathogens in community-dwelling children from Cap-Haitian, Haiti.

### Relationship between choline, betaine, DHA and diarrheal disease

Choline and DHA were associated with improved growth in Ecuador, however caregivers reported increased diarrhea for children receiving the egg intervention. We evaluated whether these nutrient biomarkers were associated with diarrheal frequency or persistence as a combined outcome. No differences were originally observed between cases and controls (37). When evaluating the association with diarrheal disease over time, no differences were observed (**Table 3**). Interestingly, DHA levels were consistently elevated in those with diarrhea at any time point (**Table 3**, column 1 relative to 2-4). No significant trends were noted based on the number of DEC identified, although choline and betaine concentrations increased in a dose dependent manner with 1 or 2 pathogens detected (**Table S7**).

**Table 3.**
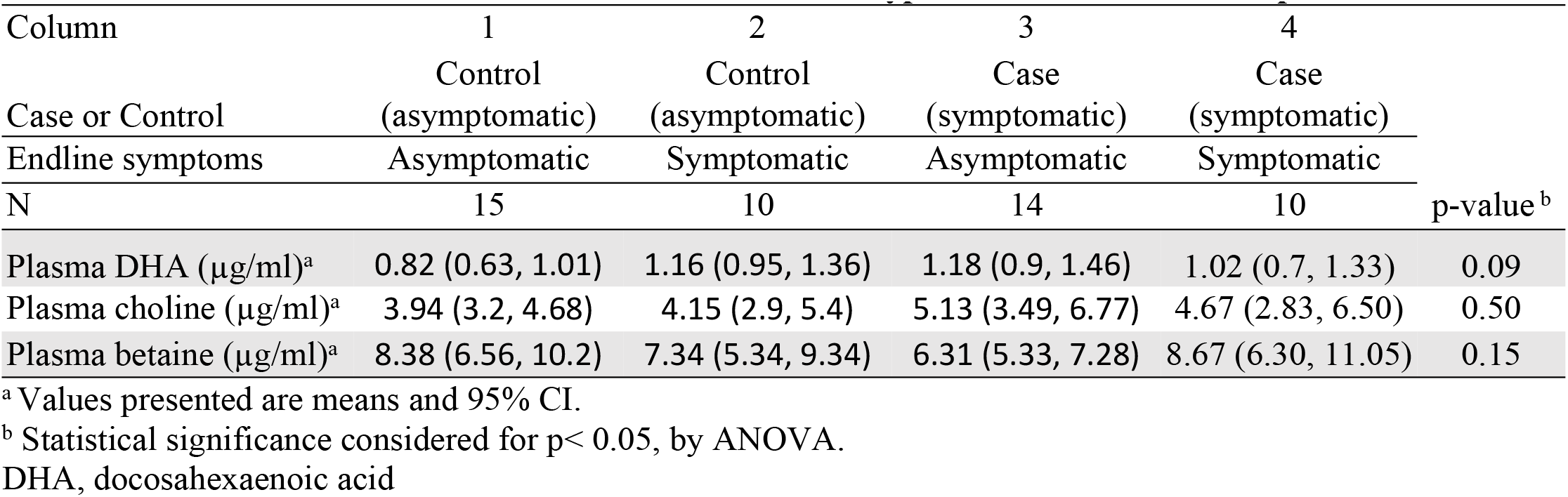
Baseline nutrient biomarkers and association with type of diarrhea at follow-up^1^.

### Relationship between pathogens and anthropometry

We evaluated the pathogen-specific association with growth faltering in Cap-Haitien to determine if distinct pathogens are associated with poor growth parameters. Univariate analysis demonstrated ETEC were significantly associated with lower mean HAZ, WAZ, and WHZ in symptomatic children at baseline (**Figure 1**). In contrast, the presence of ETEC was not associated with anthropometric measures in asymptomatic children despite similar trends. EPEC was associated with lower WHZ scores. As expected, the change in anthropometry over 1 month and or follow-up time points were not associated with significant differences (not shown).

**Figure 1.**
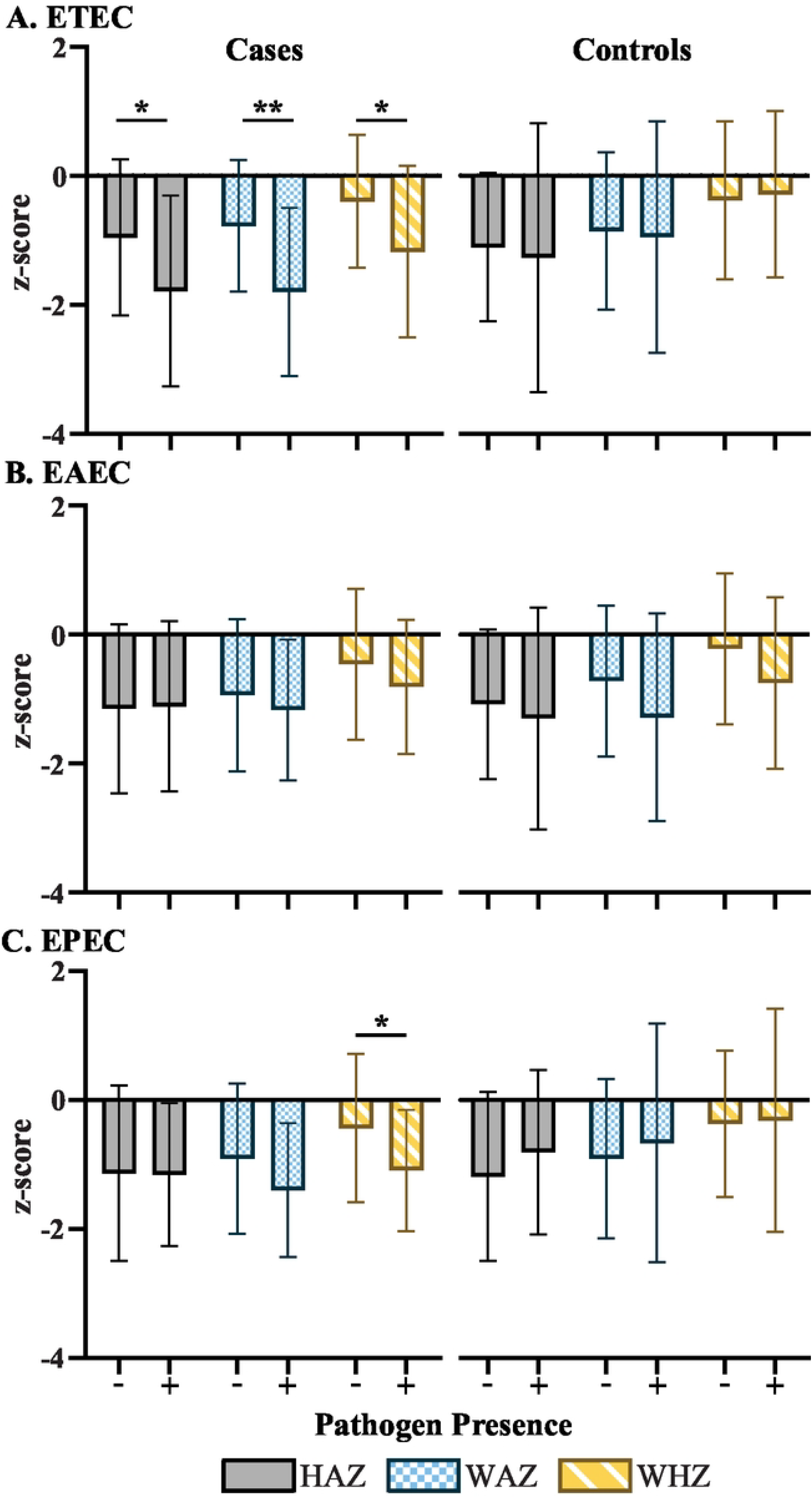
Relationship between anthropometry and DEC based on case-control status. Presence or absence of pathogenic *E. coli* is designated (+, present) and (-, absent). Student t-test compared mean z-scores based on the presence or absence of pathogens for cases (left) and controls (right). Anthropometry is designated as HAZ (Height for age z-score, solid fill), WAZ (Weight for age z-score, blue checkered fill), and WHZ (Weight for Height z-score, yellow diagonal fill). * p< 0.05 and ** p<0.001. EAEC, enteroaggregative *Escherichia coli*; EPEC, enteropathogenic *Escherichia coli*; *ETEC*, enterotoxin enterotoxigenic *Escherichia coli*.

Using multivariate linear regression, ETEC and EAEC were found significantly associated with lower WAZ and WHZ at baseline (**Table 4)**. The presence of ETEC was also significantly associated with reduced HAZ at baseline. After including the interaction term between ETEC and EAEC, the main effect of ETEC on HAZ was diminished (p > 0.15), and the interaction effect was significantly associated with lower HAZ (**Table 4**). Additionally, including the interaction term improved the proportion of variance explained by the model. The presence of symptoms was not a significant predictor of baseline anthropometric z-scores in any models.

**Table 4.** Multilinear regression models predicting anthropometry at baseline^a^.

|  | Height-age-z-score<sup>1</sup> |  |  | Weight-age-z-score |  |  | Weight-height-z-score |  |  |
| --- | --- | --- | --- | --- | --- | --- | --- | --- | --- |
|  | Coefficient B (SE) | p-value | Adjusted R<sup>2</sup> | Coefficient B (SE) | p-value | Adjusted R<sup>2</sup> | Coefficient B (SE) | p-value | Adjusted R<sup>2</sup> |
| ETEC or EAEC as sole pathogen in model |  |  |  |  |  |  |  |  |  |
| ETEC | -0.64 (0.26) | **0.014** | 0.109 | -0.69 (0.27) | **0.006** | 0.067 | -0.56 (0.24) | **0.018** | 0.081 |
| EAEC | -0.22 (0.23) | 0.33 | 0.078 | -0.49 (0.22) | **0.024** | 0.051 | -0.47 (0.20) | **0.022** | 0.079 |
| ETEC, EAEC, and ETEC*EAEC in model |  |  |  |  |  |  |  |  |  |
| ETEC*EAEC | -1.65 (0.55) | **0.003** | 0.114 | -0.72 (0.52) | 0.17 | 0.084 | 0.02 (0.51) | 0.96 | 0.056 |
| ETEC | -0.09 (0.28) | 0.75 |  |  |  |  |  |  |  |
| EAEC | 0.16 (0.23) | 0.47 |  |  |  |  |  |  |  |

<sup>a</sup>Variables included in the model based on significance in univariate analysis on anthropometry. Variables were: for HAZ, number of children in the household, animal-sourced food intake, diarrhea at baseline, sex, access to electricity, and current breastfeeding; WAZ, the number of children in the household, animal-sourced food intake, and diarrhea at baseline; for WHZ, the variables for WAZ were included plus the minimum dietary diversity and household dietary diversity scores. ETEC, enterotoxigenic *Escherichia coli*; EAEC, enteroaggregative *Escherichia coli*

## Discussion

Precise estimates of the harms associated with childhood diarrheal disease remain difficult to assess. However, these efforts are needed to prioritize prevention strategies in resource constrained environments. We quantify the burden of DEC and the association with anthropometric measures in a unique setting of Northern Haiti. We identified Haitian-specific factors that impact outcomes of diarrheal disease to inform local stakeholders on potential prevention strategies.

A high burden of diarheagenic *E. coli* exists in Cap-Haitien with current estimates closely approximating those found in other epidemiological studies. However, prior work in older school-aged children from southern Haiti suggest the pathogen-specific burden was much lower (23). Older children likely developed protection from adaptive immunity to reduce the burden of disease (43). Also, the molecular testing used in this study detects more pathogens over culture based testing used in prior work. These contrasts highlight the importance of detailed, local epidemiological studies.

Molecular diagnostics are a feasible approach in settings without established culture methodologies but present unique challenges (44). Increased pathogen detection in controls makes causal associations with diarrheal disease difficult. In comparing cases and controls, only ST-ETEC (including ST-LT ETEC) was associated with diarrheal disease, consistent with prior work in Bangladesh and elsewhere (9, 10, 42). LT-ETEC likely induces immunity after the first exposure which may not be true for ST, accounting for increased pathogenicity (9, 42). Additional virulence factors, including colonization factors, EatA, and EtpA, may further influence this risk but was not observed in our limited sample (11, 42). We observed a high prevalence of EAEC, and like ETEC, subsets of virulence factors were not associated with disease (13). Overall, the prevalence and pathogenicity of ETEC, EAEC, and tEPEC are similar to other settings. Other enteropathogens may significantly contribute to diarrhea in this population. High rates of rotavirus vaccination coverage and cholera elimination suggest these two pathogens are unlikely to significantly contribute to health outcomes (18). However, the re-emergence of cholera in Port-au-Prince may alter this presumption in future studies. COVID-19 may cause diarrheal disease, but rates in Haiti at the time of the study remained low and likely a non-significant contributor. Other context-specific SES or demographic factors may influence local epidemiology but we are unable to assess these associations at this time.

Based on prior studies, we assessed the association between plasma choline, betaine, or DHA and diarrheal disease (31, 32). DHA levels appeared higher with increased diarrheal disease, which may reflect anti-inflammatory properties of DHA. Also, a recent trial in Malawi did not show associations between DHA, choline, and growth (45), but the Malawian population differed in dietary intake, likely attenuating effects on DHA and choline. The limited sample size precludes any conclusions, but DHA and choline levels do not appear to significantly fluctuate during acute diarrheal illness.

Anthropometric measures also serve as a surrogate for poor nutrition and health outcomes (30). The association between ETEC and poor anthropometric measures has been previously observed but not reassessed in an area with high rotavirus vaccination coverage and decreasing diarrheal mortality (46, 47). EAEC was also associated with lower WHZ at baseline, supporting a potential role for this pathogen in health outcomes despite a lack of strong association with symptomatic disease (48). An interaction effect between ETEC and EAEC supports the concept that pathogens may interact synergistically, resulting in worse health outcomes for children (12, 49). A successfully licensed ETEC vaccine may therefore have benefits beyond simply reducing the adverse effects of ETEC infections alone.

The associations between anthropometry and specific pathogens have multiple explanations. First, pathogen specific virulence factors, including toxins or the presence of *eatA* or *etpA* (ETEC) or aaf genes (EAEC), may alter host-pathogen interactions and increase intestinal damage. Cultural practices may alter the risk of exposure to enteropathogens (50). Importantly, our models only explain a small portion of the variance, suggesting unmeasured confounders continue to contribute to impaired anthropometric measures. We previously identified ASFs as significant co-variates and ASF intake varies based on geography, suggesting the presence of a local context influencing growth outcomes and associations with diarrheal disease. Other possible confounders include immune status, dietary intake, and WASH practices.

Our study was not designed to determine the effect of pathogens on growth due to its case-control design, just the association between anthropometry and pathogens. Therefore, our findings must be interpreted in the setting of additional limitations of the observational study. First, we rely on care-giver reports in our survey which may introduce bias. Importantly, we were limited in our sample size due to pandemic and unanticipated financial constraints, especially when evaluating nutritional biomarkers. Since the study design does not allow us to predict the impact an episode of diarrhea has on growth, prospective studies with intensive sampling and broader pathogen detection are required in Haiti. Importantly, this study allows for refinement of future studies to focus on Haitian-specific contexts and will be tested using the framework of an on-going randomized trial (36).

Overall, our study demonstrates a high burden of DEC in Cap-Haitien, Haiti. The presence of specific DEC are associated with poor anthropometric measures and support prevention efforts targeting these pathogens. In particular, ETEC vaccines may have a significant impact on improving the health of children in Cap-Haitien. Future studies will permit estimates of the benefits related to eliminating specific enteropathogens in Cap-Haitien.

## Data Availability

All relevant data are within the manuscript and its Supporting Information files.

## Supporting Information Captions

**Table S1. Definitions of survey variables**.

**Table S2. Primers and target genes used in polymerase chain reactions**.

**Table S3. Baseline characteristics based on study completion**.

**Table S4. Numbers of respondents in Table 1**

**Table S5. Identification of multiple pathogenic E. *coli* by symptoms**.

**Table S6. Diarrheagenic *E. coli* during rainy or dry seasons**.

**Table S7. Nutritional biomarker concentrations relative to total DEC detected**.

**Database. Raw data for variables used in the analysis with variable definitions**.

## Notes

### Competing Interest Statement

The authors have declared no competing interest.

### Funding Statement

This work was funded by a grant from the Office of the Vice Chancellor for Research at Washington University (FMK, LLI) and the NIH (JND, T32 DK077653). The funders had no role in study design, data collection and analysis, decision to publish, or preparation of the manuscript.

### Author Declarations

The Washington University Institutional Review Board (ID#202007027) and the Comité National de Bioéthique in Haiti approved the study.

